# Minimally Invasive Aortic Root Surgery Without Sternotomy: Clinical and Quality-of-Life Benefits of a Totally Endoscopic Approach

**DOI:** 10.64898/2026.06.06.26354391

**Authors:** Marwan Hamiko, Saad Salamate, Ali Bayram, Florian Piekarski, Julia Rogaczewski, Kaveh Eghbalzadeh, Miriam Silaschi, Jacquline Kruse;, Ali El-Sayed Ahmad, Farhad Bakhtiary

## Abstract

**Background:** Totally endoscopic aortic root (AR) surgery via right anterior minithoracotomy (RAMT) may reduce surgical trauma and accelerate recovery compared with full sternotomy (FS). However, the approach is technically demanding due to limited access and anatomical complexity. This study compares early clinical outcomes and quality of life (QoL) after RAMT versus FS to evaluate the feasibility and safety of the totally endoscopic approach.

**Methods:** This single-center, retrospective study included 149 patients underwent AR surgery via RAMT (n=74) or FS (n=75) between January 2021 and March 2026. Patients with aortic dissection, infective endocarditis, redo surgery, concomitant procedures, or arch replacement were excluded. Operative outcomes, postoperative recovery, 30-day and 1-year mortality were analyzed. QoL was assessed using the Short Form-8 (SF-8) questionnaire.

**Results:** The median age was 60.0 years, and 79.9% of patients were male. Bentall procedure was performed in 84.6% of patients, 15.4% underwent a David procedure. Compared with FS-AR, RAMT-AR was associated with shorter median operative time (147.0 vs. 178.0 min; p<0.001), lower median chest drainage volume (650.0 vs. 850.0 mL; p<0.001), and shorter median ICU stay (24.0 vs. 25.0 h; p=0.008) and hospital stay (6.0 vs. 8.0 days; p=0.028). Overall, 30-day and 1-year mortality was 0.7%. SF-8 analysis demonstrated significantly higher physical and mental component scores in RAMT-AR patients.

**Conclusion:** In specialized centers, totally endoscopic AR surgery via RAMT is a safe and feasible minimally invasive approach associated with favorable early outcomes and a potential benefit in postoperative physical and mental QoL by reducing surgical trauma.

## Introduction

Over the past decade, minimally invasive cardiac surgery has undergone a remarkable evolution, driven by the goal of reducing surgical trauma while maintaining safety and efficacy. This progress has extended to complex aortic procedures, particularly surgery of the ascending aorta (AA) and aortic root (AR). Traditionally, these procedures have been performed via full median sternotomy (FS), which remains the gold standard in many centers.^1,2^ However, growing surgical experience and technological innovations have fostered interest in less invasive alternatives.

Partial upper sternotomy (PUS) is an established option in specialized centers, balancing adequate exposure with reduced invasiveness. Compared with FS, PUS is associated with less postoperative pain, shorter intensive care unit (ICU) and hospital stays, lower transfusion requirements, and faster mobilization, although it still involves sternotomy-related trauma.^3–8^

To further minimize invasiveness, the right anterior minithoracotomy (RAMT), initially refined for mitral and aortic valve surgery, has emerged as a promising alternative.^9–15^ Studies demonstrate superior perioperative outcomes with RAMT versus FS, including faster recovery, reduced bleeding, and improved cosmesis.^11,16–18^ In high-volume centers, RAMT has been extended to complex aortic procedures, including totally endoscopic AR replacement.

Totally endoscopic AR surgery remains technically demanding due to complex pathologies, limited exposure, challenging anatomy, and a distant operative field. These factors contribute to a steep learning curve. Nevertheless, standardized protocols and advanced technologies for minimally invasive cardiac surgery have proven effective in mitigating these challenges.^19–21^

This study compares early clinical outcomes and quality of life following totally endoscopic AR surgery via RAMT with conventional full sternotomy, evaluating its potential to redefine the standard of care in selected patients.

## Methods

### Study Population

Following approval by the institutional ethics committee of the University of Bonn (reference no. 464/22) and acquisition of written informed consent, a total of 716 adult patients were retrospectively screened between January 2021 and March 2026. After application of predefined inclusion and exclusion criteria, 149 patients undergoing elective AR surgery via RAMT or FS were included in the final analysis **(Figure 1)**, in accordance with EACTS/STS guidelines.^22^ Whenever anatomically feasible, valve-sparing root replacement (VSRR; David procedure) was performed; otherwise, patients underwent AR replacement according to the Bentall technique.

**Figure 1.**
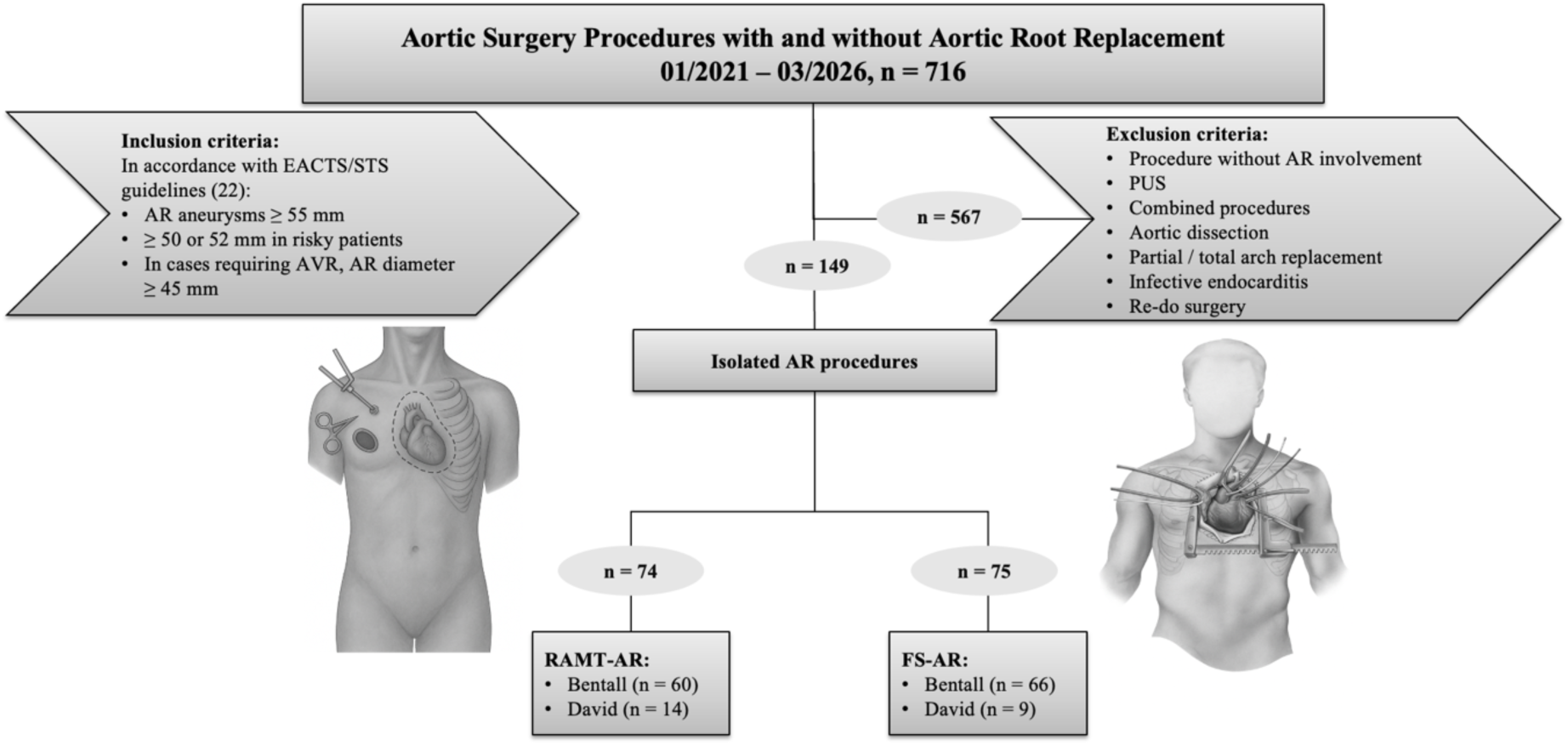
Patient selection and enrollment flow diagram. AR: aortic root; FS: full sternotomy; PUS: partial upper sternotomy; RAMT: right anterior minithoracotomy.

Baseline characteristics comprised cardiovascular risk factors, relevant comorbidities, echocardiographic and computed tomography (CT) findings, as well as New York Heart Association (NYHA) functional class. Preoperative CT imaging was routinely used to assess suitability for the minimally invasive approach, including determination of the optimal intercostal space (ICS) and cardiopulmonary bypass (CPB) cannulation strategy. Eligibility for RAMT required adequate femoral access vessels, absence of significant abdominal and thoracic aortic calcification, and a distal AA diameter ≤ 42 mm to allow safe aortic cross-clamping and adequate distal anastomotic cuff (minimum length 2 cm).

Perioperative and postoperative data were obtained from institutional electronic medical records. Preoperative operative risk was assessed using the EuroSCORE II (European System for Cardiac Operative Risk Evaluation). Primary endpoints included operative characteristics, reoperation for bleeding, and duration of ICU and hospital stay. Secondary endpoints comprised in-hospital and 30-day mortality, major adverse cardiovascular events (MACE), and aortic-related reinterventions.

### Study Groups

- **RAMT-AR:** Aortic Root Replacement via RAMT approach, n = 74
- **FS-AR:** Aortic Root Replacement via FS approach, n = 75

### Surgical Techniques

### Aortic root replacement with composite valve conduit (Bentall-Procedure)

The Bentall procedure was performed via FS or RAMT **(Figure 2A)**, depending on anatomy and surgical strategy. CPB was established via distal AA (in FS-AR patients) or femoral artery (in RAMT-AR patients) cannulation, with venous drainage through a two-stage right atrial or femoral cannula. Following aortic cross-clamping, myocardial protection was provided using antegrade Brettschneider cardioplegia (Custodiol, Dr. Franz Kohler Chemie GmbH, Bensheim, Germany); in patients with significant aortic regurgitation, cardioplegia was delivered directly into the coronary ostia after aortotomy.

**Figure 2.**
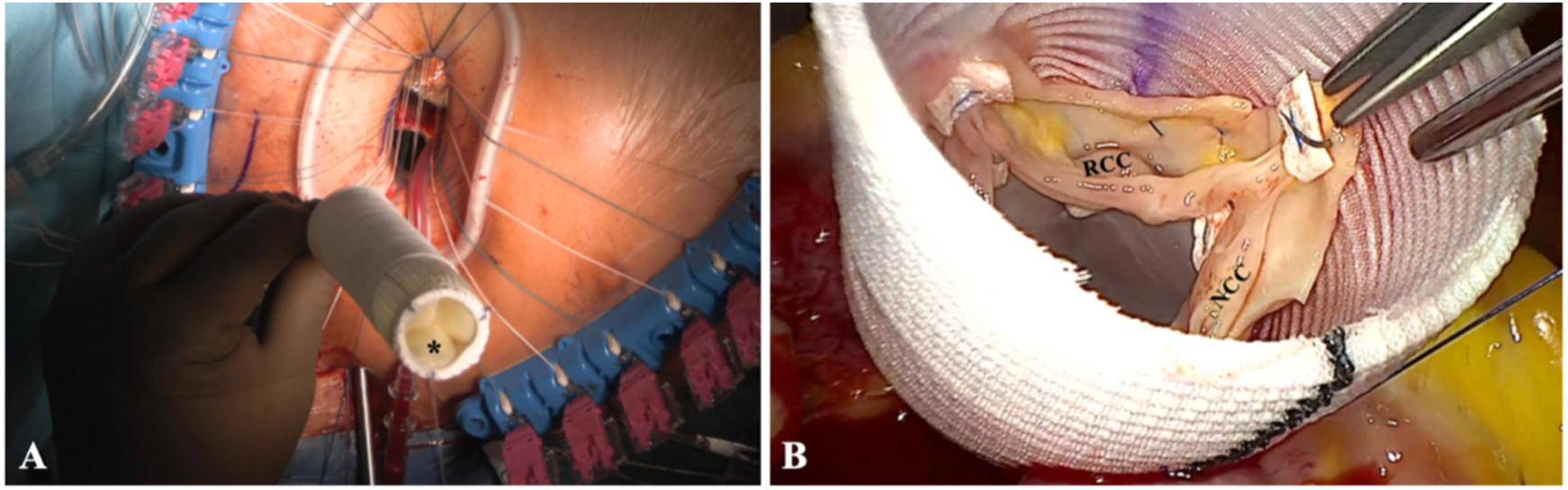
Surgical view from the 3D camera showing the RAMT-Bentall (A) using a biological conduit (**\***) and the RAMT-David (B) procedure. *NCC: non coronary cusp; RCC: right coronary cusp*.

The biological Bentall via RAMT was performed as previously described.^23^ Briefly, the AA and native aortic valve were excised, and coronary buttons mobilized. The annulus was sized, and a composite conduit was constructed by suturing a bioprosthetic valve into a vascular graft (Vascutek Gelweave, Terumo, Scotland, UK) with continuous 3-0 polypropylene during aortic cross-clamp time. In selected cases, a novel biological valved conduit (KONECT RESILIA^TM^; Edwards Lifesciences, Irvine, California, USA) was implanted according to surgeon preference and anatomical suitability. Annular sutures were placed either manually with pledgeted mattress sutures or using the RAM® device and secured with the Cor-Knot® system (LSI Solutions, Victor, NY, USA). The conduit was seated, and coronary buttons reimplanted with 5-0 polypropylene. The distal graft-to-aorta anastomosis was completed with 4-0 polypropylene.

Before unclamping, the graft and left ventricle were thoroughly de-aired under TEE guidance, and epicardial pacing wires were placed. Cannulas were removed, femoral access closed using vascular closure systems, and pleural and pericardial drains inserted. All incisions were closed according to institutional standards.

### Valve-Sparing Aortic Root Replacement (David Procedure)

VSRR using the David reimplantation technique was performed in all patients undergoing AR reconstruction. Procedures were conducted via FS or RAMT **(Figure 2B)**, based on anatomical features and surgical strategy. In RAMT cases, initial steps followed the same approach as for the Bentall procedure.

After aortotomy, the AA was transected 1–2 cm above the sinotubular junction, and the AR was mobilized to the annulus. Aneurysmal sinus tissue was excised while preserving native valve leaflets and mobilizing the coronary ostia as buttons. Three commissural stay sutures (pledgeted 4-0 polypropylene) were placed, followed by six circumferential horizontal mattress sutures (pledgeted 2-0 polyester, Ethibond, Ethicon Inc.) positioned just below the annulus and passed through the Dacron graft to secure the valve within the prosthesis. The graft was seated, and sutures tied to anchor the annulus while avoiding subvalvular stenosis.

Remaining sinus tissue was anastomosed to the graft using the commissural 4-0 polypropylene running sutures to ensure a watertight hemostatic seal. Intraoperatively, valve competence was assessed with cold saline injection. Coronary buttons were reimplanted into the graft neo-ostia with 5-0 polypropylene, and the distal graft-to-aorta anastomosis completed using 4-0 polypropylene running suture.

After thorough deairing, the aorta was declamped, patients weaned from CPB, and TEE confirmed valve competence and absence of significant regurgitation. Hemostasis was achieved, and the chest closed per institutional standards.

### Postoperative Care

All patients were admitted to the ICU postoperatively, with early extubation and transfer to the intermediate care unit (IMC) when feasible. Selected cases underwent extubation in the operating room under a fast-track protocol. Chest tube output was recorded at 1, 2, 3, 6, and 24 hours, postoperatively. According to internal policy, bleeding was considered significant if output exceeded 400 mL/h from ICU admission until 3 hours or re-thoracotomy. Collected intra- and postoperative variables included aortic cross-clamp and CPB times, ventilation duration, ICU and hospital length of stay, and in-hospital and 30-day mortality. Reoperations and MACE were also documented.

### Follow-Up and Quality of Life

Patients underwent structured follow-up at 1, 3, and 6 months, as well as 1 year after surgery. Follow-up data were obtained primarily via telephone interviews and included survival status and any aortic reinterventions. After the 1-year follow-up, patients were evaluated annually as part of the institutional follow-up program at our aortic outpatient clinic.

At 1 month after surgery, postoperative quality of life (QoL) was assessed by structured telephone interview using the 8-Item Short Form Health Survey (SF-8). The SF-8 is a validated generic instrument derived from the SF-36 questionnaire. It is designed to evaluate health-related quality of life across eight domains, including physical functioning, bodily pain, general health, vitality, social functioning, mental health, and role limitations due to physical or emotional health. Physical Component Summary (PCS) and Mental Component Summary (MCS) scores were calculated according to the standardized SF-8 scoring algorithm, with norm-based scoring referenced to the general German population, where higher scores indicate better health status.

### Statistical analysis

Continuous variables are reported as mean ± standard deviation or median with interquartile range (IQR) based on normality assessed by Q-Q plots and the Kolmogorov–Smirnov test. Categorical data were presented as absolute numbers (n) and corresponding percentages. Group comparisons used unpaired t-test or Mann–Whitney U test for continuous variables, depending on data distribution. Pearson’s chi-square or Fisher’s exact test were used for categorical variables. A p-value <0.05 was considered significant. All statistical analyses were conducted using IBM SPSS Statistics version 29 (IBM Corp., Armonk, NY, USA) and GraphPad Prism version 8.4.3 (GraphPad Software, La Jolla, CA, USA).

## Results

### Study Population

Baseline characteristics, including comorbidities and preoperative echocardiographic and CT-based parameters, are summarized in **Table 1**, with almost no significant differences between groups. The cohort was predominantly male (79.9%) with a median age of 60.0 years (IQR: 57.0-67.0) and a median EuroSCORE II of 2.3 (IQR: 1.8–3.0). Body mass index was higher in the FS-AR group (FS-AR: 28.4 vs. RAMT-AR: 26.5 kg/m^2^; p = 0.046). Preoperative echocardiography showed combined aortic valve disease in 80 (53.7%), isolated stenosis in 29 (19.5%), and isolated regurgitation in 40 (26.8%) patients. Bicuspid aortic valve was present in 53 (35.6%), left ventricular ejection fraction was preserved in 130 (87.2%), and AR diameter ranged 45–73 mm (median: 52.0 mm). Most patients were presented with NYHA class II (47.7%) or III (36.2%).

**Table 1.**
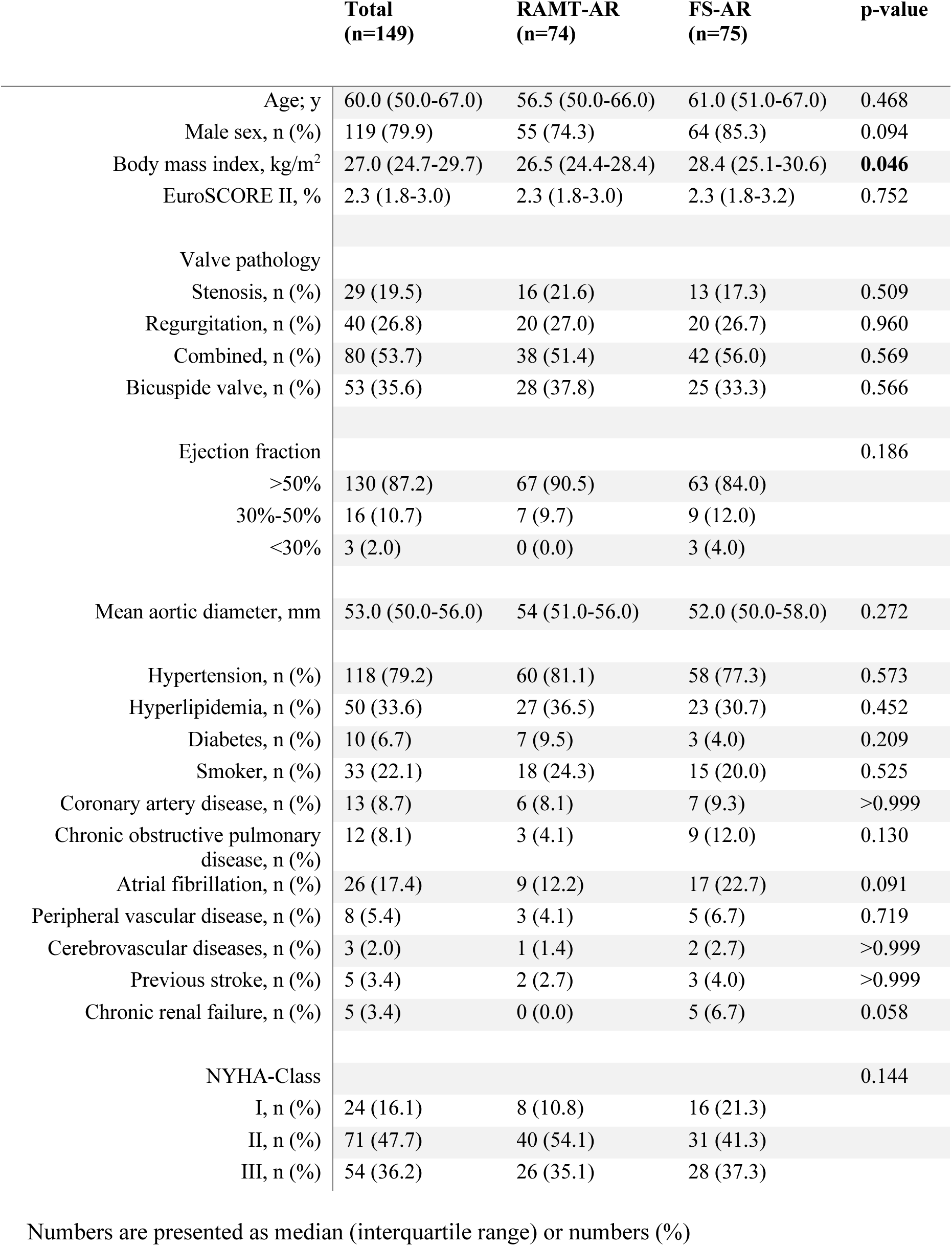
Baseline characteristics.

|  | Total<br>(n=149) | RAMT-AR<br>(n=74) | FS-AR<br>(n=75) | p-value |
| --- | --- | --- | --- | --- |
| Age; y | 60.0 (50.0-67.0) | 56.5 (50.0-66.0) | 61.0 (51.0-67.0) | 0.468 |
| Male sex, n (%) | 119 (79.9) | 55 (74.3) | 64 (85.3) | 0.094 |
| Body mass index, kg/m<sup>2</sup> | 27.0 (24.7-29.7) | 26.5 (24.4-28.4) | 28.4 (25.1-30.6) | **0.046** |
| EuroSCORE II, % | 2.3 (1.8-3.0) | 2.3 (1.8-3.0) | 2.3 (1.8-3.2) | 0.752 |
| Valve pathology |  |  |  |  |
| Stenosis, n (%) | 29 (19.5) | 16 (21.6) | 13 (17.3) | 0.509 |
| Regurgitation, n (%) | 40 (26.8) | 20 (27.0) | 20 (26.7) | 0.960 |
| Combined, n (%) | 80 (53.7) | 38 (51.4) | 42 (56.0) | 0.569 |
| Bicuspid valve, n (%) | 53 (35.6) | 28 (37.8) | 25 (33.3) | 0.566 |
| Ejection fraction |  |  |  | 0.186 |
| >50% | 130 (87.2) | 67 (90.5) | 63 (84.0) |  |
| 30%-50% | 16 (10.7) | 7 (9.7) | 9 (12.0) |  |
| <30% | 3 (2.0) | 0 (0.0) | 3 (4.0) |  |
| Mean aortic diameter, mm | 53.0 (50.0-56.0) | 54 (51.0-56.0) | 52.0 (50.0-58.0) | 0.272 |
| Hypertension, n (%) | 118 (79.2) | 60 (81.1) | 58 (77.3) | 0.573 |
| Hyperlipidemia, n (%) | 50 (33.6) | 27 (36.5) | 23 (30.7) | 0.452 |
| Diabetes, n (%) | 10 (6.7) | 7 (9.5) | 3 (4.0) | 0.209 |
| Smoker, n (%) | 33 (22.1) | 18 (24.3) | 15 (20.0) | 0.525 |
| Coronary artery disease, n (%) | 13 (8.7) | 6 (8.1) | 7 (9.3) | >0.999 |
| Chronic obstructive pulmonary disease, n (%) | 12 (8.1) | 3 (4.1) | 9 (12.0) | 0.130 |
| Atrial fibrillation, n (%) | 26 (17.4) | 9 (12.2) | 17 (22.7) | 0.091 |
| Peripheral vascular disease, n (%) | 8 (5.4) | 3 (4.1) | 5 (6.7) | 0.719 |
| Cerebrovascular diseases, n (%) | 3 (2.0) | 1 (1.4) | 2 (2.7) | >0.999 |
| Previous stroke, n (%) | 5 (3.4) | 2 (2.7) | 3 (4.0) | >0.999 |
| Chronic renal failure, n (%) | 5 (3.4) | 0 (0.0) | 5 (6.7) | 0.058 |
| NYHA-Class |  |  |  | 0.144 |
| I, n (%) | 24 (16.1) | 8 (10.8) | 16 (21.3) |  |
| II, n (%) | 71 (47.7) | 40 (54.1) | 31 (41.3) |  |
| III, n (%) | 54 (36.2) | 26 (35.1) | 28 (37.3) |  |

Numbers are presented as median (interquartile range) or numbers (%)

### Intraoperative Outcomes

Intraoperative data are summarized in **Table 2**. The Bentall procedure was performed in 126 (84.6%) patients, and the David procedure in 23 (15.4%). Among those with isolated aortic regurgitation, 17 were unsuitable for valve preservation due to advanced leaflet pathology, including fenestrations (n = 8), thickening/retraction (n = 5), or cusp sclerosis (n = 4). Biological conduits were used in 82.6% and mechanical prostheses in 1.4%, reflecting patient preference, especially in those under 60 years. Median skin to skin time was shorter in the RAMT-AR group (RAMT-AR: 147.0 vs. FS-AR: 178.0 minutes; p < 0.001). Median CPB (RAMT-AR: 111.0 vs. FS-AR: 98.0 minutes; p = 0.099) and median aortic cross-clamp (RAMT-AR: 80.5 vs. FS-AR: 76.0 minutes; p = 0.988) times were similar. None of the patients required circulatory arrest for distal anastomosis.

**Table 2.** Surgical Procedure and Intraoperative Outcome.

|  | **Total**<br>**(n=149)** | **RAMT-AR**<br>**(n=74)** | **FS-AR**<br>**(n=75)** | **p-value** |
| --- | --- | --- | --- | --- |
| Surgical Procedure |  |  |  | 0.242 |
| Bentall, n (%) | 126 (84.6) | 60 (81.1) | 66 (88.0) |  |
| David, n (%) | 23 (15.4) | 14 (18.9) | 9 (12.0) |  |
| Type of valve prosthesis |  |  |  | 0.201 |
| biological, n (%) | 124 (82.6) | 60 (81) | 64 (85) |  |
| mechanical, n (%) | 2 (1.4) | 0 (0.0) | 2 (2.7) |  |
| Duration of operation (skin to skin), min | 160.0 (145.0-190.0) | 147.0 (135.0-177.0) | 178 (150.5-207.0) | **<0.001** |
| Cardiopulmonary bypass time, min | 106.0 (90.0-125.0) | 111.0 (94.0-123.0) | 98.0 (87.0-132.0) | 0.099 |
| Aortic cross-clamp time, min | 78.0 (69.0-94.0) | 80.5 (70.0-92.0) | 76.0 (68.5-109.0) | 0.988 |

Numbers are presented as median (interquartile range) or numbers (%)

### Postoperative Outcomes

Postoperative outcomes are summarized in **Table 3**. Regrading postoperative outcome, no differences were recorded between the two study groups. In the RAMT-AR group, conversion to complete sternotomy was required in two cases (2.7%) due to uncontrollable bleeding originating from the left coronary buttons. Re-thoracotomy rate occurred in 2 patients (2.7%) in the RAMT-AR group and in 5 patients (6.7%) in the FS-AR group. Of total cohort, one (0.7%) patient experienced postoperative myocardial infarction needing extracorporeal membrane oxygenation (ECMO) due to cardiogenic shock. Acute renal failure occurred in two (1.3%) cases. One FS-AR patient (1.3%) developed disabling stroke. Postoperative delirium occurred in 9 patients (6.0%), atrial fibrillation was the most common arrhythmia (15 patients; 10.1%), and 4 patients (2.7%) required permanent pacemakers. Pneumonia occurred in 19 (12.8%), re-intubation in six patients (4.1%), including one tracheostomy (0.1%). Sternal wound infection occurred in 5 patients (3.4%), including four cases (5.3%) in the FS-AR group and one case (1.4%) in the RAMT-AR group, which required conversion to sternotomy. Cosmetic outcomes were rated excellent in RAMT-AR. In-hospital and 30-day mortality occurred in one FS-AR patient (0.7%), respectively.

**Table 3.**
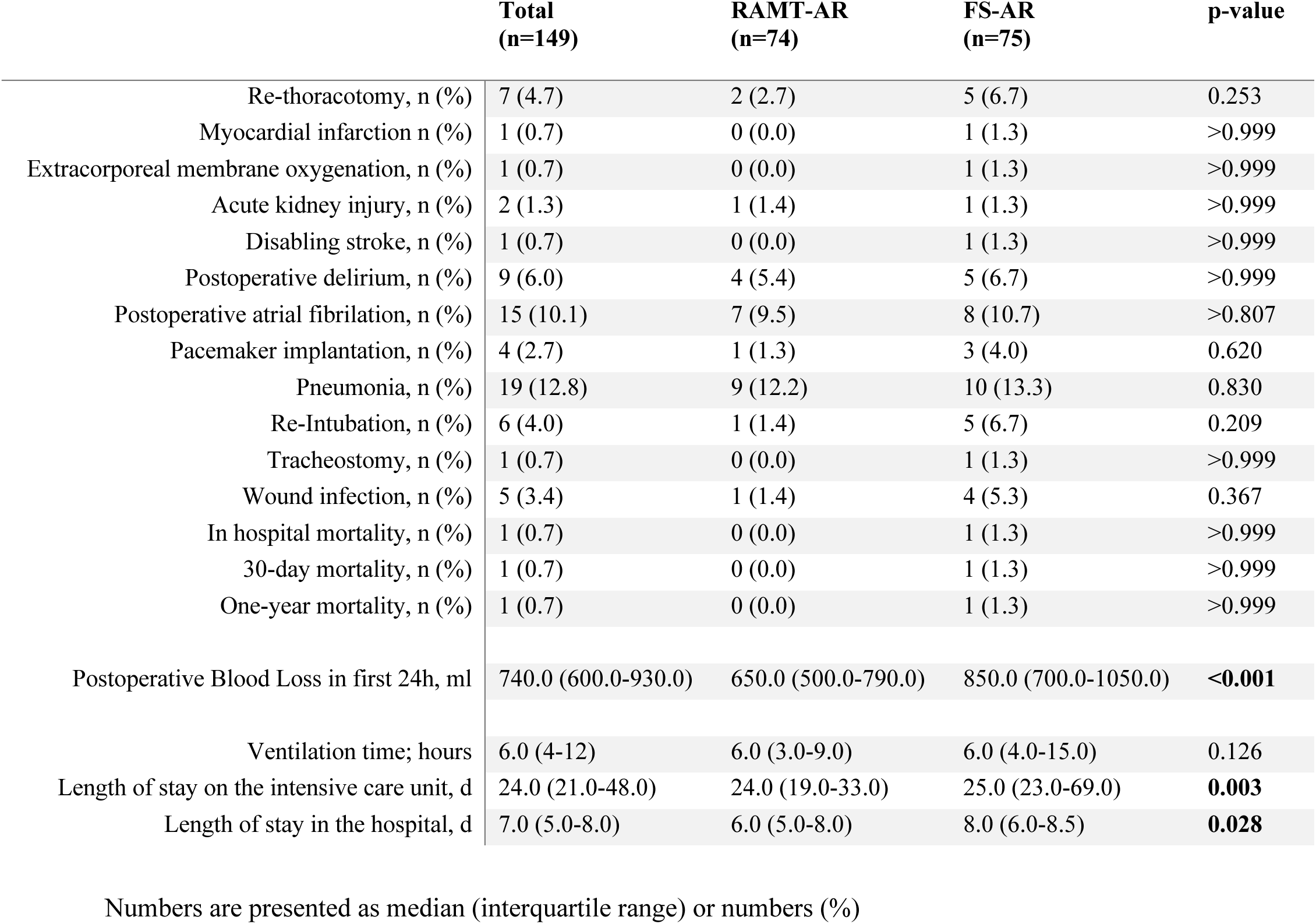
Postoperative Outcome.

Postoperative chest drainage within 24 hours was lower in RAMT-AR (RAMT-AR: 650.0 vs. FS-AR: 850.0 mL; p < 0.001). Median mechanical ventilation was comparable in both groups (RAMT-AR: 6.0 vs. FS-AR: 6.0 hours, p = 0.126). ICU stay (RAMT-AR: 24.0 vs. FS-AR: 25.0 hours; p = 0.003), and total hospital stay (6.0 vs. 8.0 days; p = 0.028) were significantly shorter in RAMT-AR.

### Follow-up

Clinical follow-up was available for 146 patients (98.0%) with a median follow-up of 12.4 months (range: 1–61 months). No cases of structural valve deterioration, late aortic-related reintervention, or mortality were observed. One year mortality was 0.7%.

### Quality of Life

The distribution of the summary scales (PCS and MCS) calculated from the SF-8 subscale scores is illustrated in **Figure 3**. RAMT-AR patients demonstrated significantly higher mean PCS (RAMT-AR: 52.1% vs. FS-AR: 46.4%; p <0.001) and MCS values (RAMT-AR: 49.6% vs. FS-AR: 42.8%; p <0.001) compared to FS-AR patients.

**Figure 3.**
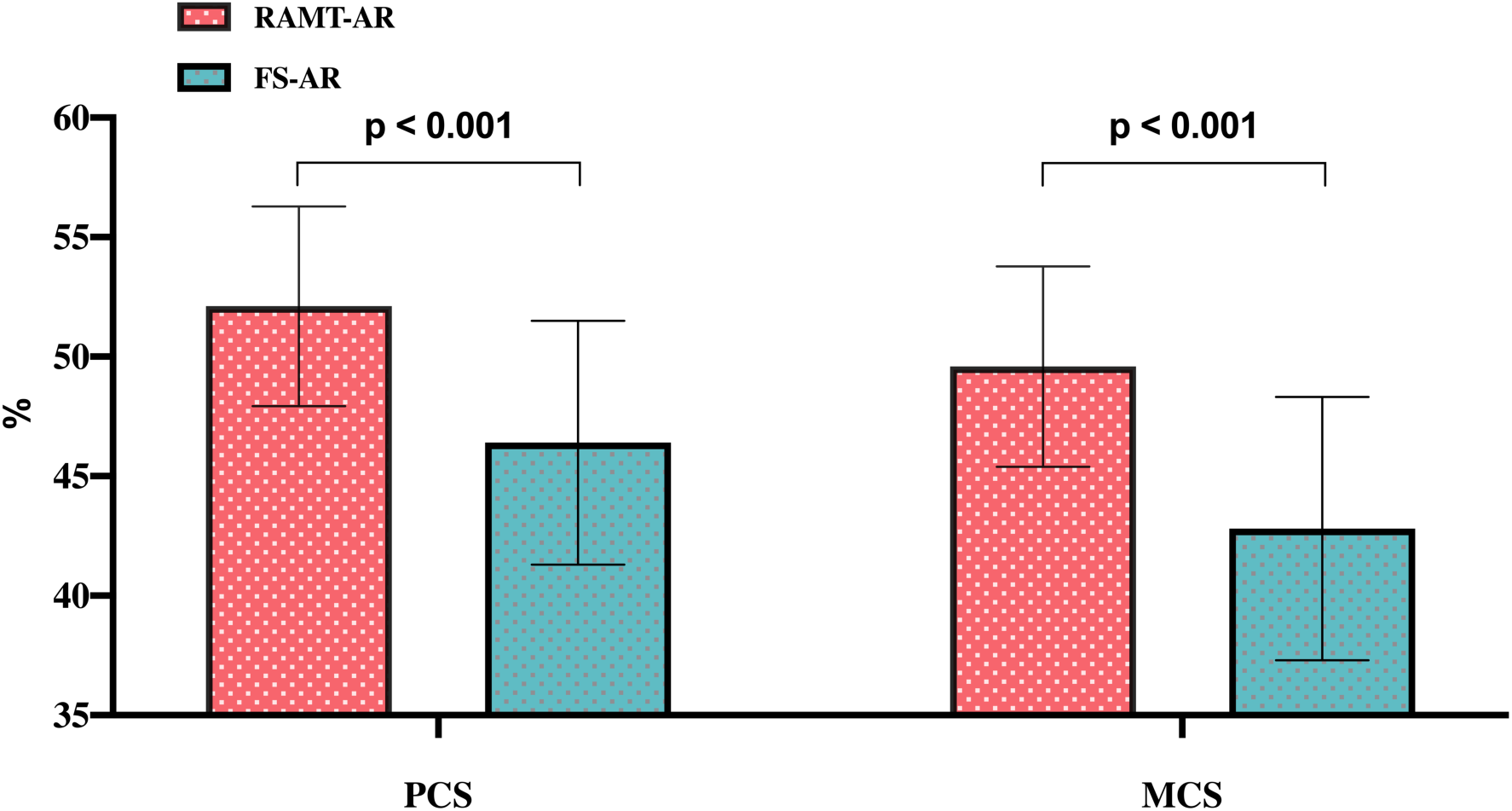
Bar plots show the mean and the standard deviation of the PCS and MCS values of RAMT-AR and FS-AR patients. *AR: aortic root, FS: full sternotomy, MCS: mental component summary, PCS: physical component summary, RAMT: right anterior minithoracotomy*.

## Discussion

Surgical management of AR pathology remains highly demanding. Median FS has traditionally served as the standard approach, providing excellent exposure and broad surgeon familiarity.^1,2^ The rise of minimally invasive cardiac surgery has challenged this paradigm, particularly in high-volume centers, by aiming to reduce surgical trauma, accelerate postoperative recovery and improve postoperative QoL.^18, 24–27^ PUS and RAMT have gained acceptance for AVR, and experience has extended their use to complex AR procedures.^23, 28^ While PUS reduces surgical trauma and maintains adequate visualization for aortic valve and proximal aortic surgery, it still involves sternal disruption and risks of impaired wound healing or sternal instability.^3–8^ Totally endoscopic RAMT may further mitigate these limitations while preserving safety and efficacy.

Our study represents one of the first direct comparisons between totally endoscopic AR surgery via RAMT and conventional FS with respect to early clinical outcome and short-term QoL. RAMT was not associated with inferior outcomes; instead, it showed significantly reduced postoperative bleeding within 24 hours, shorter operative times, and shorter ICU and hospital stays. Furthermore, the assessment of short-term QoL using the SF-8 questionnaire demonstrated significantly better physical component summary and mental component summary scores four weeks after surgery in the RAMT cohort. These findings suggest that even complex AR procedures can be safely performed using totally endoscopic approaches while simultaneously improving early patient-reported recovery and well-being.

Early experience with RAMT for AR replacement was limited to small case series. Johnson et al. reported the first RAMT Bentall procedures in seven patients, achieving satisfactory outcomes but requiring adjunctive strategies such as partial circulatory arrest and automated suture devices.^29,30^ Ji et al. expanded this to 15 patients, demonstrating feasibility without rib resection or mammary artery injury.^31^ Notably, both early studies were limited by the absence of a direct comparison with FS and were conducted using conventional instrumentation with manually constructed anastomoses. In addition, neither study incorporated an assessment of postoperative QoL.

In contrast, our technique employed high-definition 3D visualization, a 3–4 cm incision, and avoided rib retractors. Cross-clamp and CPB times were shorter than those reported by Johnson et al. or Ji et al., suggesting that optimized setup and refined expertise allow efficient and safe totally endoscopic RAMT.^29–31^ Importantly, the improved perioperative recovery observed in our RAMT cohort was paralleled by superior short-term QoL outcomes. The higher PCS scores likely reflect reduced surgical trauma, earlier mobilization, less postoperative discomfort, and faster recovery of daily physical activity. Similarly, improved MCS values may indicate lower psychological stress, greater patient satisfaction with the cosmetic result, and earlier reintegration into social activities.

Zou et al. provided the first direct comparison of minimally invasive right anterior thoracotomy (RAT) versus FS for Bentall procedures, using direct vision and rib retraction; their CPB and cross-clamp times were comparable to ours.^32^ Furthermore, Durdu et al. reported longer operative times in 31 minimally invasive RAT cases despite more valve-sparing procedures, highlighting the efficiency of our totally endoscopic approach.^33^ Consistent with these studies, ICU and hospital stays were significantly reduced following RAMT-AR procedures.^32, 33^ Mechanical ventilation times did not differ between our groups. Our institutional fast-track protocol facilitates extubation in the operating room, potentially diminishing differences between surgical approaches. Additionally, RAMT patients were sometimes transferred intubated to the ICU to avoid hypertensive surges that may predispose to postoperative bleeding.

Beyond perioperative outcomes, QoL has become an increasingly important endpoint in contemporary cardiac surgery, particularly as minimally invasive techniques continue to evolve. However, evidence specifically addressing QoL after minimally invasive AR surgery remains scarce. A recent systematic review by Hackney et al. evaluating QoL after minimally invasive AVR demonstrated that minimally invasive approaches are associated with improved early recovery, faster mobilization, shorter ICU and hospital stays, and better postoperative physical functioning compared with sternotomy.^26^ Importantly, most studies included in that review reported superior early postoperative QoL after minimally invasive surgery, although long-term differences were less consistent. The authors emphasized that minimally invasive approaches are at least non-inferior to conventional sternotomy regarding QoL and highlighted the lack of robust data for complex aortic procedures.^26^ Our findings extend these observations to totally endoscopic AR surgery and suggest that the QoL benefits previously described for minimally invasive AVR may also apply to complex AR procedures.

Similarly, the recent systematic review and meta-analysis by Marinescu et al. demonstrated that minimally invasive valve surgery was associated with significantly better short-term physical and psychological QoL outcomes compared with conventional sternotomy, particularly regarding SF-36 PCS and MCS scores. The authors attributed these improvements to reduced postoperative pain, faster rehabilitation, earlier return to normal activities, and improved body image perception.^27^ Their pooled analysis further suggested that the major QoL advantage of minimally invasive surgery occurs during the early postoperative recovery phase, whereas long-term differences tend to diminish over time. Our findings are highly consistent with these observations. By demonstrating superior SF-8 PCS and MCS scores only four weeks after surgery, our study highlights the pronounced early patient-centered benefits of totally endoscopic RAMT in AR surgery. Moreover, AR procedures are generally considered more invasive and technically demanding than isolated AVR, the observed QoL improvement is particularly noteworthy.

The improved mental health scores observed in our RAMT cohort may also reflect factors beyond reduced physical trauma. Previous studies have shown that avoidance of sternotomy may improve self-perception, reduce anxiety related to wound healing and sternal instability, and facilitate earlier recovery of independence and social functioning.^26, 27^ The totally endoscopic nature of our approach, using a limited thoracic incision without rib retractors or sternal disruption, may further enhance these effects by minimizing surgical visibility and musculoskeletal impairment.

Successful adoption of totally endoscopic AR surgery depends on surgeon experience in minimally invasive surgery, learning curve, careful patient selection, and standardized protocols. Growing experience allows inclusion of VSRR and redo procedures in the totally endoscopic program. Technological advances, including automated suture devices (RAM®, Cor-Knot®, LSI Solutions, Victor, NY, USA) and minimal-access-optimized prosthetics, enhance procedural safety and reproducibility. Minimally invasive approaches achieve clinical outcomes comparable or superior to FS, driven by reduced trauma and faster recovery. The present study additionally suggests that these perioperative advantages translate into meaningful early improvements in patient-reported physical and mental well-being. Continued development of endoscopic platforms may further improve visualization and precision, supporting broader adoption and evaluation in larger, more diverse cohorts with standardized longitudinal QoL assessment.

This study has several limitations. Its retrospective, non-randomized design introduces selection bias, as procedure choice depended on anatomical and clinical factors. The small sample size and performance by a highly experienced minimally invasive surgeon limit generalizability. The technique requires advanced proficiency in minimally invasive and conventional Bentall/David procedures, restricting its use to specialized centers. Broader adoption will require structured training and multicenter validation. Follow-up relied on telephone interviews without systematic echocardiographic or CT imaging, potentially missing detailed postoperative findings. Long-term outcomes, including valve durability and reoperation rates, warrant evaluation in prospective multicenter studies.

## Conclusion

Totally endoscopic AR surgery via RAMT is a safe and feasible alternative to conventional FS in selected patients, especially when performed in a center of excellence in minimally invasive surgery. The approach was associated with improved early perioperative outcomes, including reduced postoperative bleeding and shorter ICU and hospital stay, while also providing superior short-term physical and mental QoL recovery. These findings suggest that minimizing surgical trauma without compromising procedural safety may offer meaningful patient-centered benefits in complex AR surgery. Larger prospective studies with standardized long-term QoL assessment are warranted to further define the role of totally endoscopic approaches in contemporary aortic surgery.

## Data Availability

The data underlying this study were obtained retrospectively from clinical records and contain sensitive personal health information. Due to privacy regulations and ethical considerations, the raw data cannot be made publicly available. Researchers with a legitimate interest may request access to anonymized or aggregated data, subject to approval by the responsible ethics committee and in compliance with applicable data protection laws.

## Acknowledgments

None.

## Non-standard Abbreviations and Acronyms

AA: Ascending aorta
AR: Aortic root
AVR: Aortic valve replacement
FS: Full sternotomy
MCS: Mental Component Summary
NYHA: New York Heart Association
PCS: Physical Component Summary
PUS: Partial upper sternotomie
RAMT: Right anterior minithoracotomy
RAT: Right anterior thoracotomy
SF-8: Short Form-8 Health Survey
VSRR: Valve-sparing aortic root replacement

---

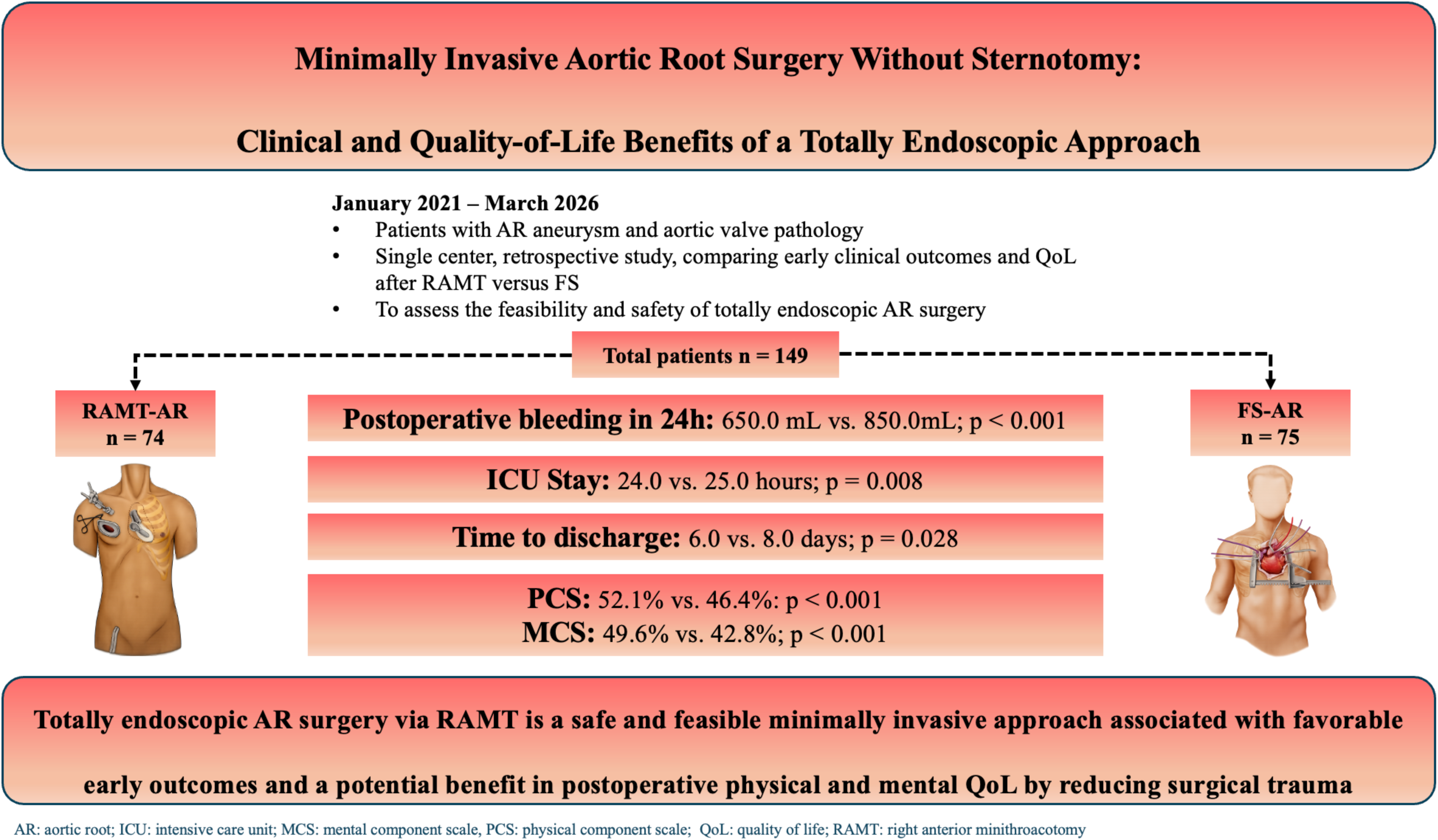

